# Measuring extracellular human brain pH and amino acid metabolism with hyperpolarized [1-^13^C]pyruvate

**DOI:** 10.1101/2023.03.23.23287579

**Authors:** Alixander S Khan, Mary A McLean, Joshua D Kaggie, Ines Horvat-Menih, Tomasz Matys, Rolf F Schulte, Matthew J Locke, Ashley Grimmer, Pascal Wodtke, Elizabeth Latimer, Amy Frary, Martin J Graves, Ferdia A Gallagher

## Abstract

Hyperpolarized carbon-13 MRI has shown promise for non-invasive assessment of the cerebral metabolism of [1-^13^C]pyruvate in both healthy volunteers and in patients. Exchange of pyruvate to lactate catalyzed by lactate dehydrogenase (LDH), and pyruvate flux to bicarbonate through pyruvate dehydrogenase (PDH), are the most widely studied reactions *in vivo*. Here we show the potential of the technique to probe other metabolic reactions in the human brain. Approximately 50 s after intravenous injection of hyperpolarized pyruvate, high flip angle pulses were used to detect cerebral ^13^C-labelled carbon dioxide (^13^CO_2_), in addition to the ^13^C-bicarbonate (H^13^CO_2_^-^) subsequently formed by carbonic anhydrase. Brain pH weighted towards the extracellular compartment was calculated from the ratio of H^13^CO_3_^-^ to ^13^CO_2_ in seven volunteers using the Henderson-Hasselbalch equation, demonstrating an average pH ± S.D. of 7.40 ± 0.02, with inter-observer reproducibility of 0.04. In addition, hyperpolarized [1-^13^C]aspartate was also detected in four of nine volunteers demonstrating irreversible pyruvate carboxylation to oxaloacetate by pyruvate carboxylase (PC), and subsequent transamination by aspartate aminotransferase (AST), with this flux being approximately 6% of that through PDH. Hyperpolarized [1-^13^C]alanine signal was also detected within the head but this was localized to muscle tissue in keeping with skeletal alanine aminotransferase (ALT) activity. The results demonstrate the potential of hyperpolarized carbon-13 MRI to assess cerebral and extracerebral [1-^13^C]pyruvate metabolism in addition to LDH and PDH activity. Non-invasive measurements of brain pH could be particularly important in assessing cerebral pathology given the wide range of disease processes that alter acid-base balance.

## 1. Introduction

Metabolic imaging with hyperpolarized (HP) ^13^C-MRI and [1-^13^C]pyruvate has been shown to non-invasively probe cellular metabolism in many tissues including the normal human brain (1,2) and cerebral tumors (3-6), with future potential applications in a range of central nervous system diseases such as stroke, multiple sclerosis, and traumatic brain injury (7,8). The technique involves intravenous injection of hyperpolarized [1-^13^C]pyruvate, which is delivered to the brain by the cerebral vasculature and transported across the blood brain barrier (BBB) by monocarboxylic acid transporters such as MCT1 into the brain parenchyma. In the cytosol, the reversible exchange of [1-^13^C]pyruvate to [1-^13^C]lactate is catalyzed by lactate dehydrogenase (LDH) and the interconversion to [1-^13^C]alanine is catalyzed by alanine aminotransferase (ALT). In comparison, pyruvate dehydrogenase (PDH) in the mitochondrial matrix catalyzes the irreversible flux to [^13^C]carbon dioxide (^13^CO_2_), which subsequently exchanges with the larger [^13^C]bicarbonate (H^13^CO_3_^-^) pool either spontaneously, or more rapidly in the presence of the enzyme carbonic anhydrase (CA) *in vivo*. Previous HP ^13^C-MRI studies in the human brain have largely focused on either [1-^13^C]lactate as the non-oxidative product of pyruvate metabolism, or [1-^13^C]bicarbonate as the oxidative equivalent. Several other metabolic pathways could potentially be investigated to provide additional biological information which have been explored here.

For example, the ratio of H^13^CO_3_^-^ to ^13^CO_2_ is pH-dependent and could therefore be used to probe acid-base balance using the Henderson-Hasselbalch equation (9):

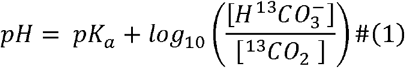

Given the critical importance of pH for the rate of many enzymatic reactions and for the maintenance of ion concentrations across the plasma membrane, normal physiological tissue pH is normally maintained within a very narrow range. In addition, cerebral pH plays a particularly important role in modulating neuronal excitation, synaptic transmission, and intercellular communication (10). Alterations in pH, particularly the development of an acidic extracellular environment, occur in many disease processes including cancer, ischemia, multiple sclerosis, and neurodegeneration (11-14). In cancer, the extracellular pH is typically lower than in healthy tissue, whilst the intracellular pH is frequently increased, with the former promoting tumor proliferation and metastasis as well as influencing the efficacy of therapy given that many chemotherapies are acidic or basic (15). Although the H^13^CO_3_^-^/^13^CO_2_ ratio can be used to probe acid-base balance, at physiological pH the ^13^CO_2_ peak is ∼36 parts per million (ppm) upfield of the H^13^CO_3_^-^ peak when assessed using ^13^C Magnetic Resonance Spectroscopy (^13^C-MRS; ∼125 and ∼161 ppm respectively) and is an order of magnitude lower, which results in the ^13^CO_2_ peak being either too small to quantify or outside the detected spectral range, or both.

HP ^13^C-MRI has been used to assess several amino acid metabolic pathways. For example, hyperpolarized [1-^13^C]alanine has been observed in the head using MR spectroscopic imaging (MRSI) following injection of hyperpolarized [1-^13^C]pyruvate in both clinical and pre-clinical studies (16,17). Although the alanine signal has been attributed to extracerebral sources, the interpretation of this has been limited by low spatial resolution and the small spectral width of the human study, which has made it difficult to identify an alanine peak separate from bicarbonate as well defining its location. Furthermore, hyperpolarized [1-^13^C]pyruvate allows for the assessment of irreversible carboxylation of pyruvate to oxaloacetate through pyruvate carboxylase (PC), and subsequent transamination to aspartate by aspartate aminotransferase (AST), which can be detected as hyperpolarized [1-^13^C]aspartate. This has been previously demonstrated in the murine liver (17) and the healthy human brain using non-hyperpolarized ^13^C-labelled glucose (18). An age-dependence of PC activity as detected using hyperpolarized [1-^13^C]pyruvate metabolism has also been reported with lower levels of labelled aspartate in older individuals (19).

This study investigated the potential of HP ^13^C-MRI to measure: cerebral pH using unlocalized MRS; [1-^13^C]alanine using MRSI and spectrally-selective spiral MRI; and hyperpolarized [1-^13^C]aspartate with slice-selective MRS. Using these approaches, we show the potential to measure pH non-invasively, as well as estimating cerebral PC flux relative to PDH flux from the relative signal strength of aspartate to bicarbonate in each volunteer.

## 2. Methods

### 2.1 Hyperpolarized ^13^C-MRI

Hyperpolarized ^13^C-MRI was performed using a 3 T MRI system (MR750; GE Healthcare, Waukesha, WI, USA) and a birdcage transmit/receive head coil utilizing quadrature detection for ^13^C, and linear detection for ^1^H (Rapid Biomedical, Rimpar, Germany). The study was performed with the approval of a local research ethics committee (MISSION-MIMS; REC: 15/EE/0255) and with the informed consent of all volunteers. The clinical fluid path containing [1-^13^C]pyruvate was assembled under aseptic conditions using a validated process described previously (1). Prior to HP ^13^C-MRI imaging, T_1_W 3D proton images were taken using a spoiled gradient recalled echo sequence. The predicted frequency of ^13^C-pyruvate was calculated from the frequency of water in this imaging series. Eddy current settings (20) were adjusted to the gyromagnetic ratio of ^13^C followed by Bloch-Siegert transmit power and frequency calibration. Mean ^13^C-pyruvate polarization was measured as 16.6% (range 6.1%-29.7%) from the 13 doses injected.

### 2.2 Imaging parameters

13 volunteers were injected with 0.4 mL/kg of ∼250 mM [1-^13^C]pyruvate up to a maximum of 40 mL at a rate of 5 mL/s. ^13^C-MRSI data from 10 of these healthy volunteers has been reported previously (21) but has been reanalyzed to investigate the distribution of ^13^C-labelled alanine and the spectral data was used to investigate aspartate levels, and to measure pH. Imaging was acquired 22-27 s after injection of ^13^C-pyruvate using the following parameters: field-of-view [FoV] = 20 cm, matrix = 10 × 10, TR = 323 ms, flip angle = 8–10°, bandwidth = 5000 Hz, 544 points, scan duration = 20 s, with 61 transients, 5 slices, 2 cm thick. Following MRSI, both unlocalized spectra and high flip angle slice-localized spectra were acquired with a sufficiently large bandwidth to measure low intensity metabolites including: ^13^CO_2_ for the estimation of global brain pH (TR = 1 s, flip angle = 45°, rectangular pulse width 0.25 ms, 15000 Hz, 4096 points, beginning 44-49 s after the start of the pyruvate injection) and ^13^C-aspartate (TR = 1 s, flip angle = 90°, 15000 Hz, 4096 points, 3-5 slices). Where both acquisitions were performed, this was followed by multiple unlocalized MRS transients to fully exhaust the hyperpolarized magnetization allowing the remaining natural abundance of ^13^C in fat to be identified separately from the hyperpolarized ^13^C peaks.

In three volunteers, high-resolution spiral imaging with metabolite-specific spectral-spatial (SPSP) (22) excitation was used to obtain dynamic measurements (nominal pixel size 6×6 mm, TR = 1 s, flip angles = 15° for pyruvate and 40° for other metabolites, 62,500 Hz, 3034 points, 3 slices, 3 cm thick, beginning 10 s after injection). This was followed by slice-localized MRS, beginning 76 s after injection. Table 1 details the acquisition scheme used for each volunteer.

**Table 1.** HP ^13^C acquisition schemes used for the 13 healthy human volunteers. A variety of MR Spectroscopic Imaging (MRSI) and Spectral-Spatial (SPSP) schemes were used to acquire the data followed by MRS in both slice selective and unlocalized forms.

| Volunteer | Acquisition scheme | Age Range | Gender | Polarization (%) |
| --- | --- | --- | --- | --- |
| 1 | 2D MRSI<br>Unlocalized MRS | 21-25 | M | 15.1 |
| 2 | 2D MRSI<br>Unlocalized MRS | 26-30 | M | 6.1 |
| 3 | 2D MRSI<br>Unlocalized MRS | 66-70 | M | 6.1 |
| 4 | 2D MRSI<br>Unlocalized MRS<br>5 slice selective MRS<br>Unlocalized MRS | 41-45 | F | 13.4 |
| 5 | 2D MRSI<br>Unlocalized MRS<br>5 slice selective MRS<br>Unlocalized MRS | 35-40 | F | 12.7 |
| 6 | 2D MRSI<br>Unlocalized MRS<br>5 slice selective MRS<br>Unlocalized MRS | 26-30 | F | 19.3 |
| 7 | 2D MRSI<br>Unlocalized MRS<br>5 slice selective MRS<br>Unlocalized MRS | 16-20 | M | 22.8 |
| 8 | 2D MRSI<br>Unlocalized MRS<br>5 slice selective MRS<br>Unlocalized MRS | 26-30 | F | 18.7 |
| 9 | 2D MRSI<br>Unlocalized MRS<br>5 slice selective MRS<br>Unlocalized MRS | 31-35 | M | 13.4 |
| 10 | 3D MRSI<br>Slice selective MRS | 36-40 | M | 13.4 |
| 11 | SPSP<br>3 slice selective MRS | 66-70 | M | 22.8 |
| 12 | SPSP<br>3 slice selective MRS | 26-30 | M | 29.7 |
| 13 | SPSP<br>3 slice selective MRS | 26-30 | F | 22.0 |

### 2.3 Data analysis

Spectra were analyzed in MATLAB (The Mathworks, Natick, MA) for the quantification of the ^13^C-bicarbonate, ^13^CO_2_, and ^13^C-aspartate peaks. Zero and first order phase correction was applied to the spectra with the same first order correction applied with each MRS acquisition scheme used: unlocalized = -0.07 rads/ppm; 3 slice selective = -0.298 rads/ppm; and 5 slice selective = -0.339 rads/ppm). Quantification of ^13^C-bicarbonate (161 ppm) and ^13^CO_2_ (∼125 ppm) peaks was performed by two independent users using baseline adjusted peak heights. In both cases, the nearest peak-free segment of the baseline either side of the peak was set to zero. The frequency profile of the excitation pulse was measured to make corrections for the measured peak heights for ^13^CO_2_ and H^13^CO_3_^-^ based on the frequency profile of the excitation pulse (Supplementary Methods). This led to a decrease in the relative ratio of H^13^CO_3_^-^/^13^CO_2_ of 0.86, and a decrease in the measured pH by 0.07 pH units. The pH was calculated using Equation 1, assuming a p*K*_a_ of 6.17 (9). Repeatability between observers was measured using the Bland-Altman test (23) and the coefficient of variation.

From the slice-selective MRS data, ^13^C-pyruvate, ^13^C-bicarbonate, and ^13^C-aspartate peaks were measured using the peak heights relative to baseline from the central slice within the brain. The signal-to-noise ratio (SNR) for the metabolites was calculated, with an aspartate SNR of 2.5 set as the limit for detection. Metabolite quantification from MRS data was again performed by two users to assess repeatability of the measurements.

The OXSA (Oxford Spectroscopy Analysis) toolbox implementation of AMARES peak fitting (24,25) was applied to MRSI to quantify levels of ^13^C-alanine, ^13^C-pyruvate, ^13^C-lactate, and ^13^C-bicarbonate within each voxel. In one subject (volunteer 12), high-resolution spiral images of ^13^C-alanine were acquired and compared to the other metabolites.

## 3. Results

An unlocalized ^13^C spectrum from volunteer 9 is shown in Fig. 1, in which both ^13^CO_2_ and H^13^CO_3_^-^ were detected. A ^13^CO_2_ peak was identified at 125.6 ppm and a bicarbonate at 161 ppm, with no detectable metabolite after 8 transient pulses decreased the hyperpolarized signal, with the remaining signal at ∼128 ppm arising from natural abundant ^13^C present within lipids.

**Figure 1.**
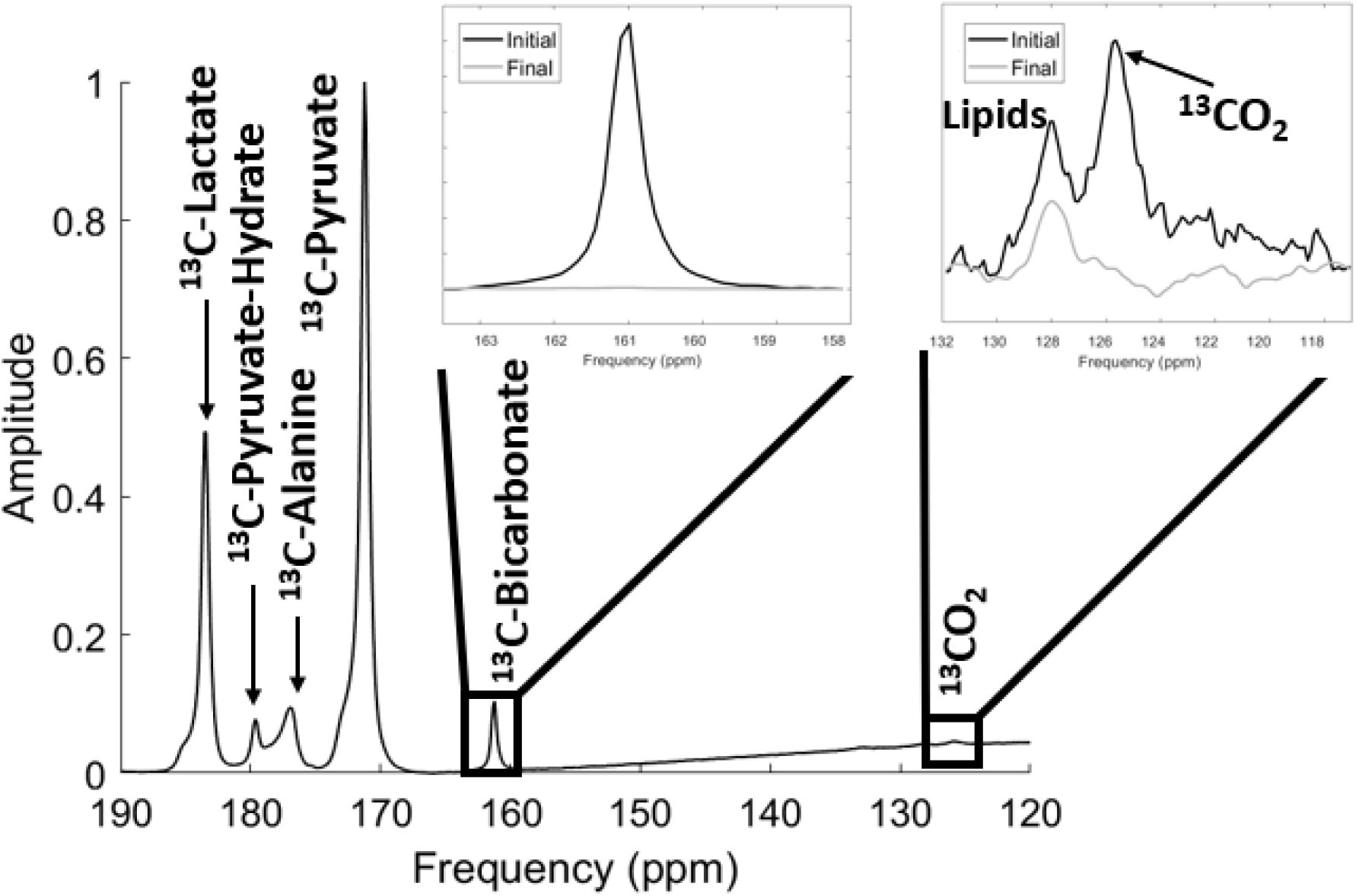
Unlocalized spectrum from subject 8 acquired using MRSI. The ^13^C labelled metabolites are shown: ^13^C-pyruvate, ^13^C-lactate, ^13^C-alanine, ^13^C-pyruvate-hydrate, ^13^C-bicarbonate, and ^13^CO_2_. The spectrum is enlarged to clearly show the ^13^C-bicarbonate and ^13^CO_2_ signals in the initial spectrum (black) and the natural abundance lipid signal after the decay in polarized signal in the final scan (grey), following 8 pulses.

Unlocalized MRS was acquired in 9 volunteers following MRSI acquisition. Fig. 2 shows the real phased spectrum for each subject from 117-132 ppm, where the ^13^CO_2_ peak can be identified at 125 ppm. A ^13^CO_2_ peak was successfully measured in 7 of the 9 volunteers (78%) with a lipid peak visible in most cases also. The decay of the ^13^CO_2_ in the final spectra, following repeated MRS pulses to exhaust the hyperpolarized signal, in comparison to the first spectra is shown, leaving only the residual natural abundance ^13^C-lipid peak remaining.

**Figure 2.**
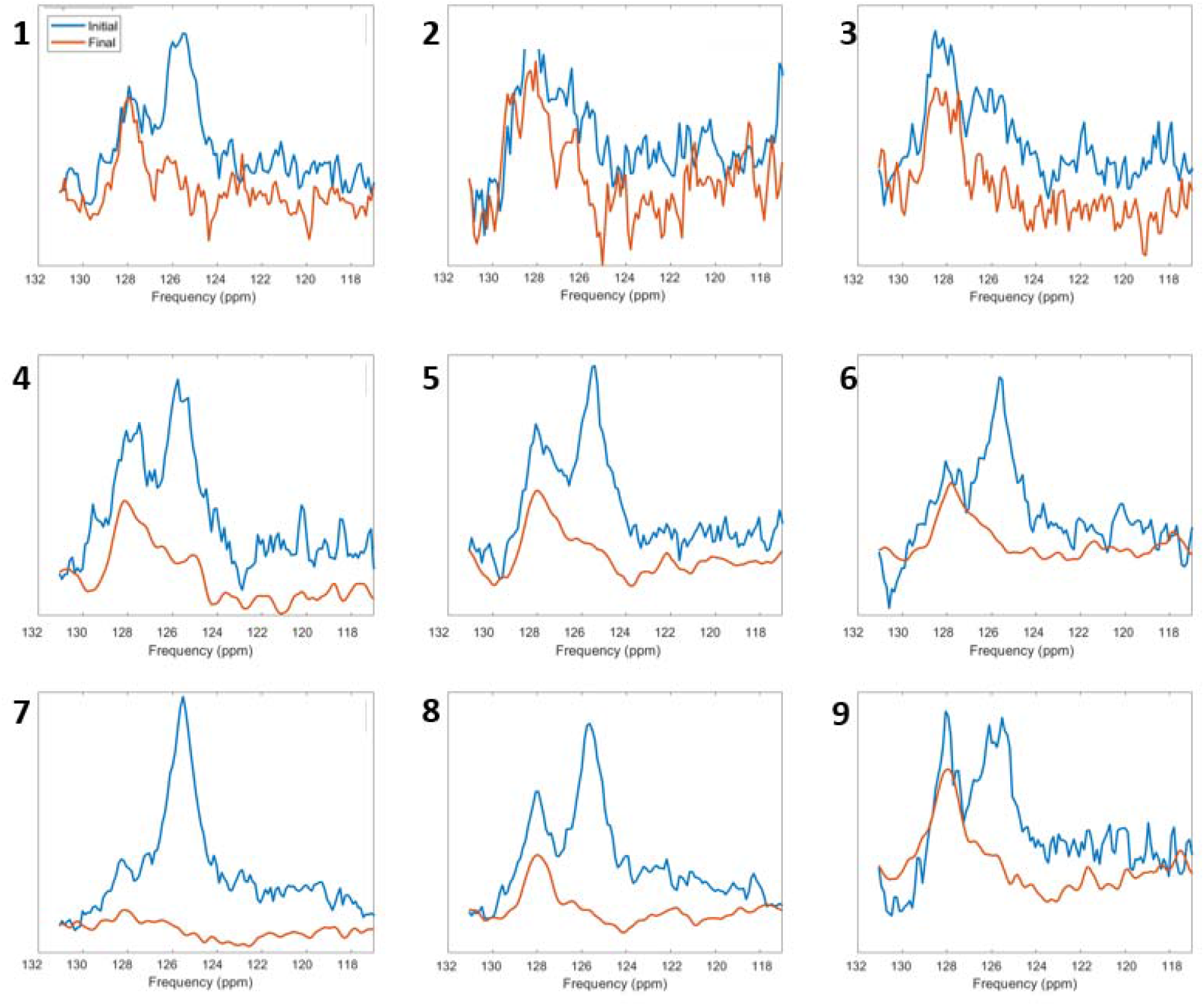
^13^C spectra from 9 volunteers with the initial (blue) and final (red) spectra shown. The hyperpolarized ^13^CO_2_ peak at 125 ppm can be seen in 7 out of the 9 cases but had decayed by the final spectra. The peak at 128 ppm reflects non-hyperpolarized naturally abundant ^13^C signal from tissue lipids.

pH was calculated and is shown in Table 2 with a mean pH ± S.D. of 7.40 ± 0.02. Repeatability between the two observers was found to be 0.04 pH units with a coefficient of variation of 0.89%. pH was not measured in volunteers 2 and 3 due to the inability to distinguish ^13^CO_2_ signal above the noise floor.

**Table 2.** pH measurements in 9 subjects that underwent unlocalized MRS. pH was measured using equation 1 by two observers independently. Peak heights were corrected for the excitation pulse frequency profile which resulted in a change of -0.07 pH units. Brain pH was estimated in 7 of the 9 cases (78%).

| Volunteer | pH measured by observer 1 | pH measured by observer 2 | Mean pH |
| --- | --- | --- | --- |
| 1 | 7.38 | 7.46 | 7.42 |
| 2 | N/A | N/A | N/A |
| 3 | N/A | N/A | N/A |
| 4 | 7.37 | 7.35 | 7.36 |
| 5 | 7.41 | 7.37 | 7.39 |
| 6 | 7.39 | 7.45 | 7.42 |
| 7 | 7.40 | 7.42 | 7.41 |
| 8 | 7.42 | 7.4 | 7.41 |
| 9 | 7.37 | 7.43 | 7.40 |
| Mean | 7.39 ± 0.02 | 7.41 ± 0.04 | 7.40 ± 0.02 |

The formation of hyperpolarized ^13^C-alanine within the head was analyzed using ^13^C-MRSI (Fig. 3a). The imaging showed marked spatial variation of the alanine signal across the head with high levels found outside the brain, particularly within the temporalis muscles bilaterally, with no significant alanine signal arising from the brain parenchyma. Higher resolution SPSP ^13^C-MRI was also performed (Fig. 3b), again confirming that muscle is the likely predominant source of the alanine signal detected.

**Figure 3.**
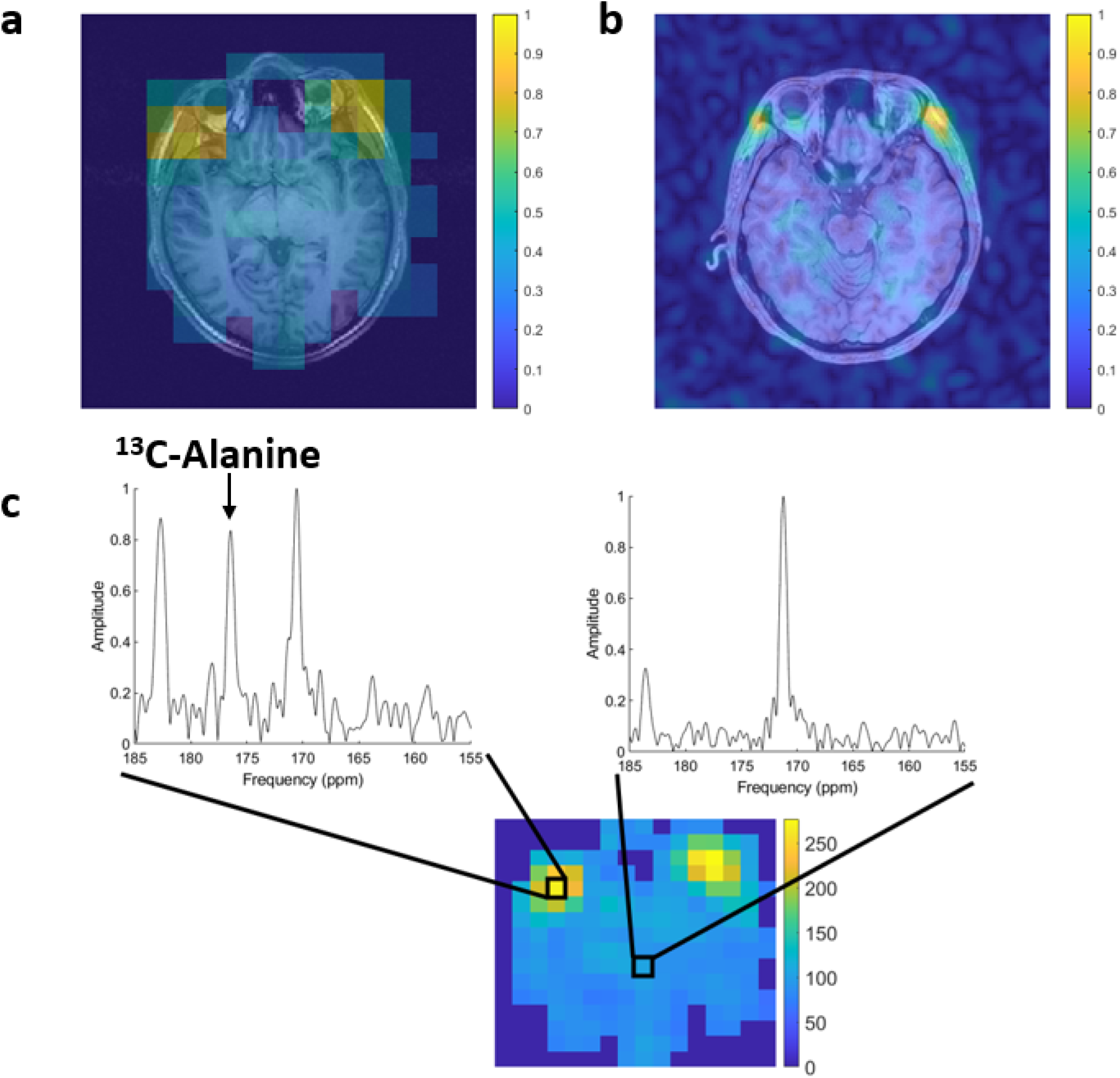
(a) ^13^C-MRSI from volunteer 2 and (b) SPSP from volunteer 11 are shown overlaid onto the proton imaging demonstrating the location of the hyperpolarized ^13^C-alanine signal, which is centred on the temporalis muscles bilaterally. (c) MRSI analysis of two voxels from volunteer 2 with different levels of hyperpolarized ^13^C-alanine signal taken from the extracerebral muscles (left) and from the brain parenchyma (right).

Slice-selective spectra were analyzed by two observers to investigate the presence of aspartate. In 4 out of the 9 volunteers an [1-^13^C]aspartate SNR greater than 2.5, the threshold for detectability, was measured with a mean SNR of 4.6 across the two observers. Table 3 lists the SNR for these cases and the time from injection to measurement for comparison. Figures 4a-c shows the spectra from three volunteers of different ages, with aspartate seen in two out of the three examples. Figure 4b shows the one case where [4-^13^C]aspartate was also identified albeit with an SNR of only 2.4. Figure 4d shows the relation between ^13^C-aspartate and ^13^C-bicarbonate SNRs in the four cases where aspartate SNR was above the detection threshold (shown in blue). A linear fit of y = 0.063x + 1.49 was found for these cases (R^2^ = 0.73) suggesting a PC/PDH flux ratio of approximately 6%.

**Table 3.** SNR measurements of hyperpolarized ^13^C-pyruvate, ^13^C-bicarbonate, and ^13^C-aspartate measured from ^13^C-MRS on a central slice through the brain. Volunteers 11-13 were imaged with a 3 slice MRS acquisition approach, whereas volunteers 5-10 underwent a 5 slice acquisition. A SNR threshold of 2.5 was set for aspartate detection.

| Volunteer | $^{13}\text{C}$-pyruvate SNR | $^{13}\text{C}$-bicarbonate SNR | $^{13}\text{C}$-aspartate SNR | Time from injection to acquisition (s) |
| --- | --- | --- | --- | --- |
| 5 | 120.4 | 32.4 | 2.4 | 52 |
| 6 | 100.7 | 28.4 | 1.9 | 52 |
| 7 | 272.6 | 76.4 | 2.4 | 52 |
| 8 | 320.8 | 57.2 | 3.8 | 52 |
| 9 | 97.5 | 20.5 | 3.1 | 52 |
| 10 | 389.1 | 51.0 | 2.4 | 32 |
| 11 | 213.8 | 44.7 | 4.6 | 76 |
| 12 | 262.4 | 77.0 | 7.1 | 76 |
| 13 | 62.4 | 13.7 | 2.3 | 92 |

**Figure 4.**
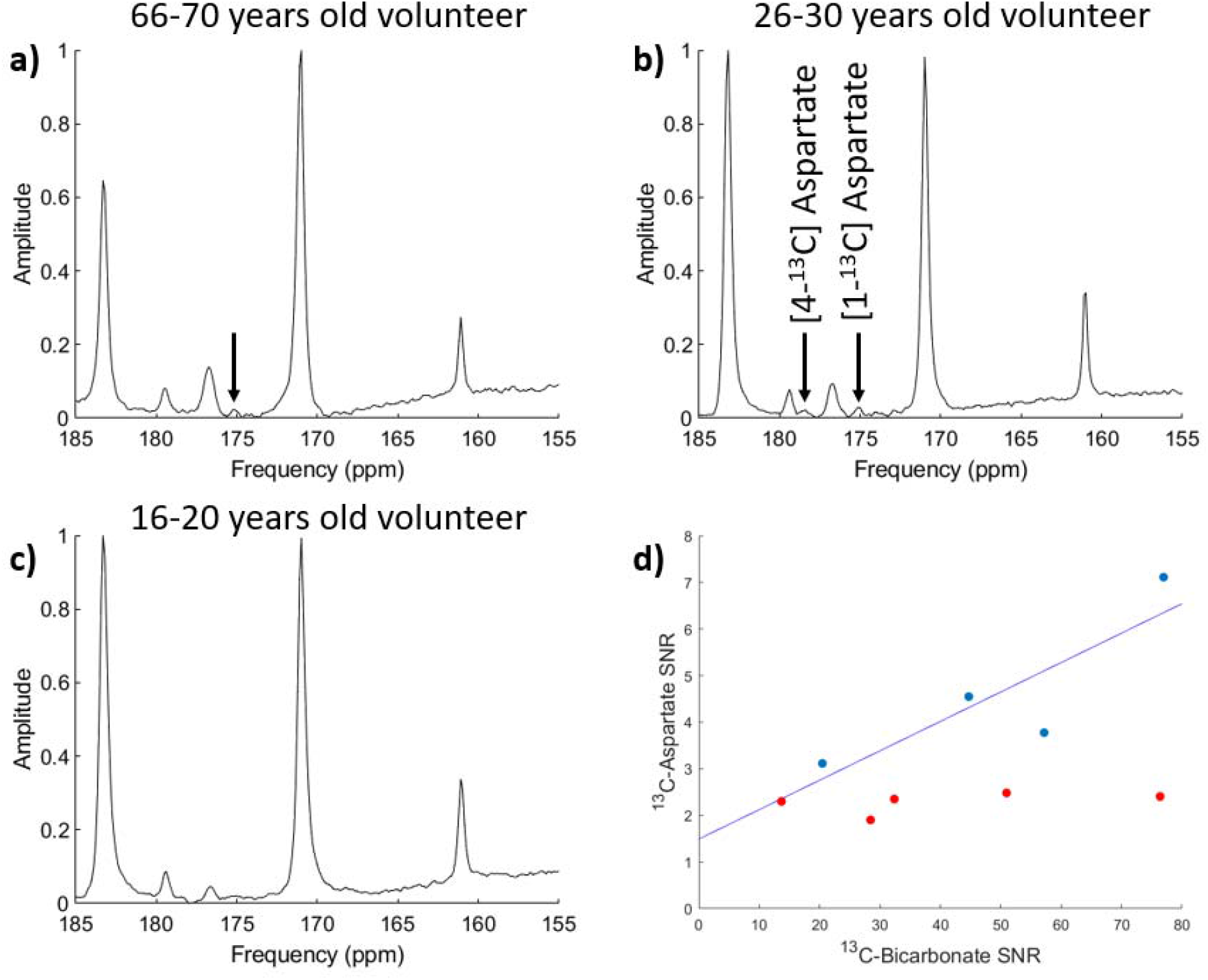
(a) ^13^C spectra from a volunteer in the age range of 66-70 years old (volunteer 11), where [1-^13^C]aspartate was detected (aspartate SNR = 4.6). (b) ^13^C spectra from a volunteer in the age range of 26-30 years old (volunteer 12) where both [1-^13^C]aspartate and [4-^13^C]aspartate were also detected (aspartate SNR = 7.1) (c) ^13^C spectra from a volunteer in the age range of 16-20 years old (volunteer 7) where aspartate was not detected above the noise floor (aspartate SNR = 2.4). (d) Correlation between ^13^C-aspartate and ^13^C-bicarbonate SNR for all 9 cases with a linear fit applied to the 4 cases (blue) where aspartate was detected above the SNR threshold of 2.5. Cases where aspartate is below the 2.5 SNR limit are shown in red.

## 4. Discussion

HP ^13^C-MRI has been widely used to probe oxidative and non-oxidative metabolism in the brain in both health and disease, through the detection of ^13^C-bicarbonate and ^13^C-lactate respectively, following the injection of hyperpolarized ^13^C-pyruvate (1-3). Here we show how injected ^13^C-pyruvate can be used to measure tissue pH and study amino acid metabolic pathways in addition. Given that the technique has now been translated into human brain imaging, these findings could have important implications for studying new cerebral biology, as well as providing novel approaches to stratify disease and response to therapy.

### 4.1 Brain pH

Many pathological conditions are associated with alterations in central nervous system pH such as ischemia, demyelination, and cancer (26-28). However, there is currently no routine clinical tool for imaging tissue pH in humans. Measuring the ratio of tissue ^13^C-labelled bicarbonate to carbon dioxide allows the endogenous buffering system to be harnessed to probe pH: not only does this allow a rapid and direct measure of tissue acid-base balance, but given the high concentrations of these molecules *in vivo*, it also mitigates against the probe acting as a buffer and therefore altering the pH it is measuring (9). Furthermore, as the approach is reliant on a ratio, it is independent of probe delivery to the tissue, assuming that the signal from both molecules is detectable above the noise floor and that they reach chemical equilibrium within the timescale of this detection. Bicarbonate and carbon dioxide are rapidly interconverted under the action of the ubiquitous enzyme carbonic anhydrase (CA), with cell membrane-bound isoforms of this enzyme such as carbonic anhydrase 9 (CAIX) having a role in maintaining an acidic extracellular pH in many tumors (29). We have previously shown that the rapid exchange between HCO_3_^-^ and CO_2_ can be exploited to measure pH with HP ^13^C-MRI, following the administration of exogenous hyperpolarized ^13^C-bicarbonate and using the ratio of the two molecules together with the known p*K*a (30): the resulting measurement is weighted towards the extracellular compartment (pH_e_). This approach has been used to measure pH in preclinical tumor models and in the rodent brain (9,31). In comparison, injected hyperpolarized ^13^C-pyruvate may also result in the intracellular formation of ^13^CO_2_ due to oxidative decarboxylation of pyruvate within the mitochondria, which subsequently forms H^13^CO_3_ ^-^ catalyzed by intracellular isoforms of CA. The ^13^CO_2_ can diffuse out of the cell down its concentration gradient and H^13^CO_3_^-^ may be transported into the extracellular space, where extracellular and membrane-bound isoforms of CA (such as CAIX and CAXII) will catalyze further exchange. The relatively long timescale (44-49 s) after the injection of ^13^C-pyruvate in these experiments before the measurement of pH, enables a significant proportion of the labelled ^13^CO_2_ and/or H^13^CO_3_^-^ to diffuse or be transported into the extracellular-extravascular space as well as the vascular compartment. Hyperpolarized ^13^C-pyruvate has been used to measure acid-base balance in the heart which is thought to be weighted towards the intracellular space (pH_i_): PDH activity is particularly high in the heart (32) which has allowed this approach to be used to measure cardiac pH with values ranging between 7.1-7.3 (33-36). Here we use hyperpolarized ^13^C-pyruvate to measure human brain pH for the first time.

Despite the low SNR of the hyperpolarized ^13^CO_2_ following hyperpolarized ^13^C-pyruvate injection, signal from the dissolved gas could be identified in 78% of volunteers enabling pH to be calculated. The average cerebral pH measured by the two readers in the seven volunteers was 7.40 ± 0.02 showing a good agreement across subjects. The interobserver reproducibility was 0.04 pH units and a coefficient of variation of 0.89% demonstrated the robustness of these measurements. These results compare favorably to other methods that have attempted to measure extracellular brain pH (37). Invasive methods to probe brain pH involve the insertion of microelectrodes into the brain with electrochemical sensors to measure extracellular pH (38): whilst offering a very direct measurement, the invasive method cannot be used for routine clinical applications and the insertion of the probe may lead to inflammation which may alter pH. Non-invasive MR spectroscopic methods have been used to determine pH *in vivo* such as ^31^P-MRS. The chemical shift of the inorganic phosphate peak (P_i_) is dependent on intracellular pH with a p*K*a of 6.73 and can be measured relative to phosphocreatine (PCr) and alpha-ATP which are pH-independent (39). However, even though the peaks are visible without the need for exogenous compounds, ^31^P-MRS has not yet been adopted into clinical practice, which in part may be explained by the larger and earlier changes in extracellular pH with the onset of disease compared with those seen with intracellular pH, and therefore non-invasive measurements of extracellular pH are of particular interest clinically. Recent advances in HP ^13^C-MRI have led to new hyperpolarized pH-responsive probes such as [1,5-^13^C_2_]zymonic acid (40) which offer the potential to measure extravascular/extracellular and intravascular pH with a suitably long T_1_ for *in vivo* applications. Investigations into the application of a similar hyperpolarized probe in preclinical studies has shown the potential for pH mapping in the rodent kidney (41). This demonstrates good agreement with electrode pH measurements and offers the ability for high resolution pH mapping but this has yet to be translated into the clinic.

The p*K*a for the exchange reaction between CO_2_ and bicarbonate is approximately 6.17, which is close to the physiological pH range, resulting in the technique being most sensitive to changes in the pH range of greatest biological interest. Hyperpolarized ^13^C-bicarbonate has not yet been translated into the clinic, which is partly due to the short relaxation time of its hyperpolarized signal (42). In comparison, HP [1-^13^C]pyruvate has been widely used as part of clinical studies to assess alterations in tissue lactate labelling: measurements of extracellular pH using this approach could provide important additional biological information in conjunction with the metabolic information provided by measuring the exchange of pyruvate to lactate and irreversible flux to bicarbonate. Previous studies have utilized ^13^C-pyruvate to map pH in the rat heart with recorded cardiac pH values in the range of 7.12-7.30 (33-35). Measurements of human cardiac pH using the same approach have been reported in a single volunteer undergoing a non-selective radiofrequency pulse acquisition to measure whole heart pH (36): the recorded pH of 7.1 was weighted more towards the intracellular pH which may be explained by the very high cardiac PDH activity and the early timepoint of acquisition after injection (<29 s). The brain also demonstrates high expression of the enzyme PDH compared to most other tissues, which has enabled the measurement of cerebral pH here (7).

Several technical requirements are needed to enable ^13^CO_2_ concentration to be accurately measured within the brain, and therefore enable pH measurement. As the ^13^CO_2_ frequency is considerably lower than the other main products of [1-^13^C]pyruvate metabolism, a large bandwidth is needed to simultaneously observe both ^13^CO_2_ and ^13^C-bicarbonate: we used a non-slice-selective rectangular pulse with a duration of 0.25 ms, giving a bandwidth of 4000 Hz or 125 ppm, which included the entire range of metabolites between ^13^C-lactate and ^13^CO_2_. Slice selection with spectral-spatial excitation alternating between the frequencies of ^13^CO_2_ and ^13^C-bicarbonate would be an alternative approach; however, the low SNR of the ^13^CO_2_ signal limits the spatial resolution. By using an unlocalized spectrum, a larger volume of the brain could be interrogated increasing the sensitivity for ^13^CO_2_ detection given that its concentration is an order of magnitude smaller than H^13^CO_3_^-^. To verify that the peak was derived from hyperpolarized ^13^C, and not from naturally abundant signal, a series of spectra were acquired with a large excitation flip angle to fully deplete the magnetization. In all volunteers the ^13^CO_2_ peak had decayed in the final transient with only the peaks attributed to the naturally abundant ^13^C in lipid remaining. In the subjects where no ^13^CO_2_ peak was visualized above the noise floor, there was low initial polarization (6.1% in both cases) which was less than half the average polarization for the other subjects (15.2%), emphasizing that pH measurements can be achieved in the context of a reasonable level of polarization. Although we increased the bandwidth of the excitation pulse to measure pH, the ^13^CO_2_ peak was still just outside the optimal frequency window. To account for this, the relative peak height of ^13^CO_2_ and bicarbonate was corrected using an empirically determined adjustment. The frequency could be optimally centered on ^13^CO_2_ to maximize the signal of this difficult to measure species, or midway between it and bicarbonate to minimize uncertainty in the relative weighting of the two molecules.

The measured cerebral pH here using hyperpolarized ^13^C-pyruvate closely resembled the expected extracellular value. Although a large fraction of brain volume is composed of the intracellular compartment (43), the extracellular bicarbonate concentration is more than double that of the intracellular compartment which is partly controlled by high astrocytic activity which modulates bicarbonate levels in the extracellular space and may result in the measured hyperpolarized pH being weighted towards the extracellular space (44). Moreover, the extracellular weighting of the pH will also be facilitated by the rapid diffusion of CO_2_ and transportation of HCO_3_^-^ over the relatively long timescale of the experiment (44-49 s) and may also undergo vascular washout in this timescale and therefore this pH may in addition be weighted to towards the venous blood pH. The spectral resolution of ^31^P-MRS can be used to separate the intracellular and extracellular components of the P_i_ signal, and using this approach the intracellular cerebral pH has been recorded as 7.0 with an extracellular pH of 7.4 (37), the latter aligning well with the results we have obtained with HP ^13^C-MRI here. Invasive pH measurements of rat brains using microelectrode pH sensors to measure extracellular pH have estimated values ranging between 7.21-7.27 (38). We have previously shown that the rodent brain pH measured using ^13^C-MRS and injected hyperpolarized ^13^C-bicarbonate was 7.20 ± 0.08 (31) which supports the hypothesis that this is likely to represent a contribution from both the intracellular and extracellular compartments. An assumption is that the p*K*_a_ value used in equation 1 is the same in brain tissue compared to other organs, but the close alignment between the results here and the ^31^P-MRS and invasive measurements of pH suggest that this assumption is likely to be correct. The measured pH is not only weighted towards the extracellular pool but is also likely to be heavily weighted towards grey matter: previous studies have demonstrated that bicarbonate signal is highest in grey matter, reflecting its high perfusion and metabolic activity (1,2), and therefore the measured pH here is also likely to reflect signal from grey matter.

### 4.2 Aspartate

In four of the nine cases investigated with slice-selective MRS, it was possible to detect [1-^13^C]aspartate at an SNR level greater than 2.5. In one case, [4-^13^C]aspartate was also clearly seen (Fig 4b). Although a previous report has suggested age-related trends in ^13^C-aspartate signal (19), our data did not demonstrate this as is illustrated in Fig. 4, which shows the spectra from three different volunteers of different ages: ^13^C-aspartate levels were high in three young volunteers and undetectable in five further young volunteers despite being detectable in a volunteer in the age range of 66-70 years-old.

Hyperpolarized [1-^13^C]pyruvate has a relatively limited capacity to probe tricarboxylic acid (TCA cycle) metabolism due to the loss of the ^13^C label as ^13^CO_2_, catalyzed by pyruvate dehydrogenase, when pyruvate enters the cycle. Alternative labelling strategies include using hyperpolarized [2-^13^C]pyruvate, which has been shown to label downstream [5-^13^C]glutamate as an indirect biomarker of TCA activity (45). However, often the chemical shifts, relaxation times, and J-coupling properties of the TCA cycle intermediates are less favourable for *in vivo* imaging studies (46). [1-^13^C]pyruvate also allows an indirect assessment of pyruvate carboxylase (PC) activity if hyperpolarized ^13^C-aspartate can be detected due to the irreversible carboxylation of pyruvate to oxaloacetate by pyruvate carboxylase (PC), and subsequent transamination by aspartate aminotransferase (AST) as has been demonstrated in the mouse liver (17) and the healthy human brain (19). Age-dependence of PC activity has been suggested, with lower levels of aspartate being detected at older ages (47,48). Since bicarbonate signal is a measure of the irreversible flux through PDH activity, the ratio of PC flux compared to PDH flux can be estimated from the relative signal strength of ^13^C-aspartate to ^13^C-bicarbonate. Previous investigations using [2-^13^C]glucose at thermal polarization have found that the PC flux accounted for 6% of the rate of glutamine synthesis (18), and here we have confirmed this using HP ^13^C-pyruvate for the first time.

The ^13^C-aspartate/^13^C-bicarbonate ratio was used here as a quantitative biomarker to estimate the PC/PDH ratio *in vivo*. Similar ^13^C-aspartate/^13^C-bicarbonate ratios were seen in subjects where spectra were acquired 76 s after injection using a spectrally selective spiral acquisition approach, and in those where the MRS acquisition began at 52 s following multi-slice MRSI. The ^13^C-aspartate SNR was compared to that of ^13^C-bicarbonate for all subjects, including those who fell below the aspartate SNR threshold of 2.5. Overall, the spectral SNR was not the limiting factor in most of the cases where aspartate was not detected, which agrees with an earlier study where [1-^13^C]aspartate was detected in only five of nine subjects (19), so the reason for the lack of detection may be due to fundamental biological differences between individuals rather than technical limitations. Ideally, aspartate would be measured in the spectra acquired for ^13^CO_2_ measurement, but it was found that the magnetic field homogeneity in these whole-head spectra was insufficient to resolve the aspartate peaks from the larger nearby ^13^C-alanine and ^13^C-pyruvate-hydrate signals. A linear fit acquired from those subjects where aspartate was detected suggested flux through PC is approximately ∼6% of the PDH flux.

The [1-^13^C]aspartate and [4-^13^C]aspartate peaks exist at the same ppm as [1,5-^13^C_2_]zymonic acid at physiological pH. Since some zymonic acid may contaminate [1-^13^C]pyruvic acid, or be generated from pyruvate in aqueous solution, there is potential for zymonic acid to be mischaracterised as aspartate, but dynamic phantom data using hyperpolarized ^13^C pyruvate did not show the presence of zymonic acid in this study [Supplementary Materials]. Within the healthy brain, the expression of PC is glia-specific (49), and its role is to ensure TCA activity by compensating for losses of α-ketoglutarate that occur through transmitter release of glutamate and GABA in neighboring neurons (50). However, brain damage or other pathological changes result in metabolic alterations, with the relative PC/PDH flux altering depending on the specific metabolic requirements. This has been shown in rat models of traumatic brain injury (TBI) (51), with metabolism altering to favor PC activity, potentially suggesting different metabolic priorities of the brain following damage. In two cases (volunteers 8 and 9), pH and aspartate were measured in the same imaging session using one pharmacy kit following MRSI imaging, showing how this could be a simple addition to existing protocols. Therefore, the PC/PDH ratio may be a useful non-invasive HP ^13^C-MRI biomarker of differential pyruvate metabolism in the brain when aspartate can be detected, and future studies could explore its role in cerebral pathology.

### 4.3 Alanine

Previous human HP ^13^C-MRI studies have demonstrated hyperpolarized ^13^C-alanine within the pancreas and muscle (52), including potential signal arising from temporalis muscles within the head (53). However, in the latter study it has been difficult to discriminate between ^13^C-alanine and ^13^C-bicarbonate due to spectral overlap on Echo-Planar Spectroscopic Imaging (EPSI) sequences which results in the bicarbonate peak being folded into the spectrum close to the alanine peak, rendering them difficult to distinguish (16). To overcome this, in this study we investigated the presence of alanine within the head in healthy human volunteers using full-bandwidth MRSI. Fig. 3a shows the ^13^C-alanine signal is extremely high in voxels localized to the temporalis muscles, and in some cases nearly equivalent to the ^13^C-lactate signal and much larger than the ^13^C-bicarbonate signal. In comparison there is no detectable alanine signal within the brain parenchyma as expected since alanine aminotransferase (ALT) is expressed at low levels (54) within the brain compared to the liver and muscle. Localization of the hyperpolarized ^13^C-alanine within the muscle was further confirmed using the high resolution SPSP imaging as shown in Fig. 3b. This high muscle metabolism of ^13^C-pyruvate on HP ^13^C-MRI is analogous to the high uptake of radiolabelled ^18^F-fluorodeoxyglucose on Positron Emission Tomography (^18^F-FDG PET) in muscle that is seen when movement has occurred during tracer uptake and image acquisition (55) and may also relate to muscle activity in these experiments.

## Conclusion

Hyperpolarized ^13^C-MRS offers a non-invasive method to probe tissue metabolism that has been extensively used preclinically to image metabolic processes within the brain, and more recently has been translated into human imaging. To date, the method has largely focused on the exchange of injected ^13^C-pyruvate into ^13^C-lactate, and in this study, we show that the technique can be used to provide additional biological information which may have important clinical applications. Using both unlocalized and slice-selective spectroscopy, we non-invasively measured brain pH and estimated the ratio of pyruvate flux through pyruvate carboxylase and pyruvate dehydrogenase. This study also investigated the localization of hyperpolarized ^13^C-alanine and verified that the signal originates from muscle outside the brain. These measurements have the potential for providing important additional information that could be useful metrics for improving diagnosis and stratification of disease, as well as early measurement of success response to treatment response across a range of disorders.

## Data Availability

All data produced in the present study are available upon reasonable request to the authors

## Acknowledgements

We have funding from Cancer Research UK (CRUK; C19212/A27150; C19212/A16628), the Lundbeck Foundation, the Multiple Sclerosis Society (2015–35), the National Institute of Health Research (NIHR) Cambridge Biomedical Research center (BRC-1215–20014), CRUK Cambridge center, the Cambridge Experimental Cancer Medicine center, the Evelyn Trust, Addenbrooke’s Charitable Trust, and the NIHR/Wellcome Trust Cambridge Clinical Research Facility.

## CRediT authorship contribution statement

**Alixander S Khan:** Conceptualization, Data curation, Software, Writing – review & editing. **Mary A McLean:** Conceptualization, Data curation, Software, Writing – review & editing. **Josh D Kaggie:** Conceptualization, Data curation, Writing – review & editing. **Ines Horvat-Menih:** Data curation, Writing – review & editing. **Tomasz Matys:** Conceptualization, Writing-review & editing. **Rolf F Schulte**: Software, Writing – review & editing. **Matthew J Locke**: Conceptualization, Data curation, Writing – review & editing. **Ashley Grimmer**: Data curation, Writing – review & editing. **Pascal Wodtke**: Conceptualization, Writing-review & editing. **Elizabeth Latimer**: Data curation, Writing – review & editing. **Amy Frary**: Data curation, Writing – review & editing. **Martin J Graves**: Conceptualization, Writing – review & editing. **Ferdia A Gallagher:** Conceptualization, Data curation, Software, Writing – review & editing.

