## Supplementary Material for "Measuring extracellular human brain pH and amino acid metabolism with hyperpolarized [1-^13^C]pyruvate"

**Supplementary methods**

RF excitation pulse frequency characterization

The frequency profile of a 252 μs hard pulse was characterized using a frequency sweep on a phantom. Spectra were acquired at the center frequency and iterated at 100 Hz offsets, moving the center frequency from the phantom frequency. SNR values were calculated at each offset in order to find the change in SNR in relation to frequency offset.

In addition, the magnitude of the magnetization in the XY plane was simulated in Matlab by directly solving the Bloch equation in the rotating frame.

Dynamic Phantom data

Unlocalized MRS was performed in a phantom of hyperpolarized ^13^C pyruvate (spectral width 5000 Hz, 2048 points, flip 3°, 320 averages).

**Supplementary results**

RF excitation pulse frequency characteristics

^13^C-bicarbonate is detected at 161 ppm, 331 Hz away from the transmit frequency (pyruvate at 171 ppm). This results in a measured intensity of 95 % relative to ^13^C-pyruvate (Suppl. Fig. 1A).^13^CO_2_ is detected at 125 ppm, 1478 Hz away from pyruvate, which results in a measured relative intensity of 82 %. The relative intensity of ^13^CO2/^13^C-bicarbonate would therefore be 0.82/0.95 = 0.86. Solving the Bloch equation for a rectangular pulse of this duration confirms sensitivity at the ^13^CO_2_ offset to be 82% of maximum (Fig. 1B).

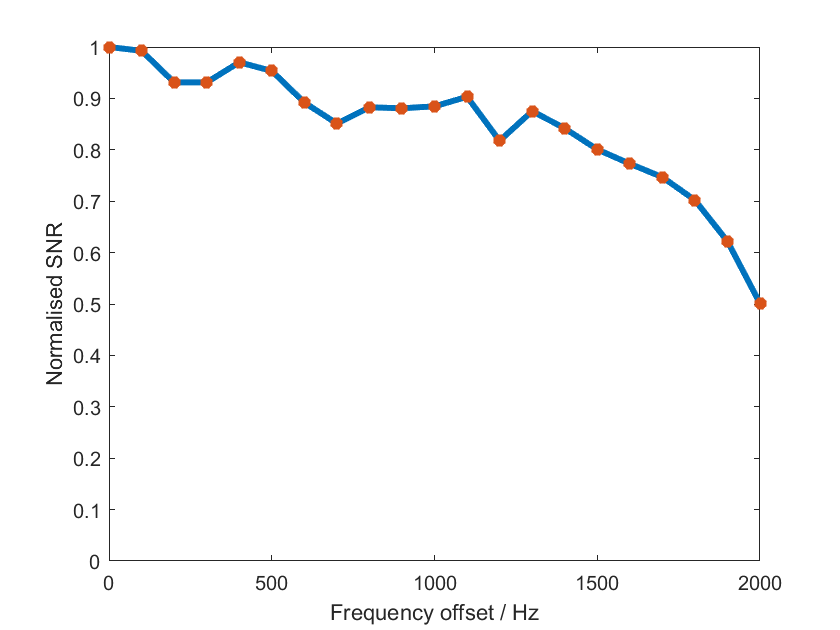

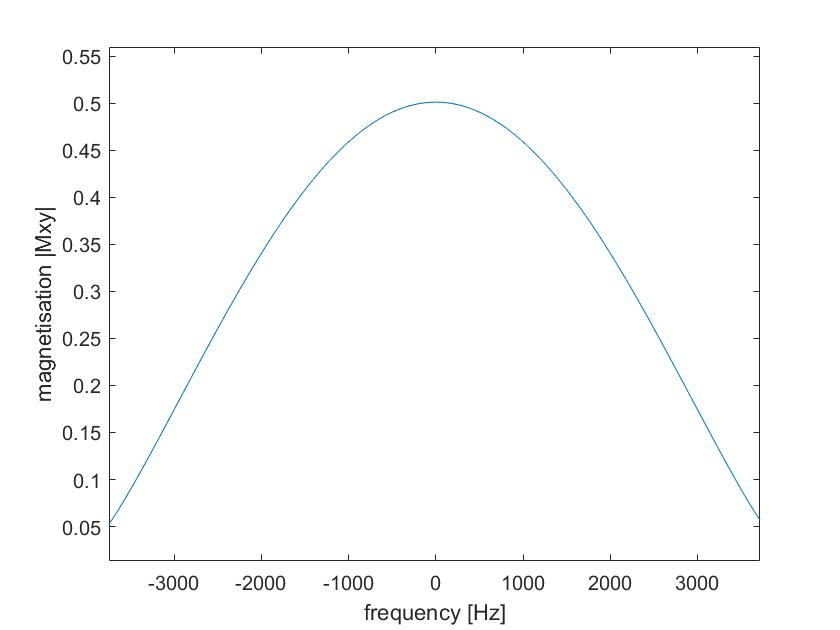

Supplementary figure 1: (A) Measured frequency profile of the RF excitation pulse from the center frequency (set to that for pyruvate) up to 2000 Hz (B) Calculated frequency profile of a 252 μs rectangular pulse obtained by solving the Bloch equation in the rotating frame.

Hyperpolarized ^13^Cpyruvate in dynamic phantom data

In the dynamic phantom data only ^13^C-pyruvate (171 ppm) and ^13^C-pyruvate hydrate (176 ppm) can be seen showing that zymonic acid is not contaminating the samples.

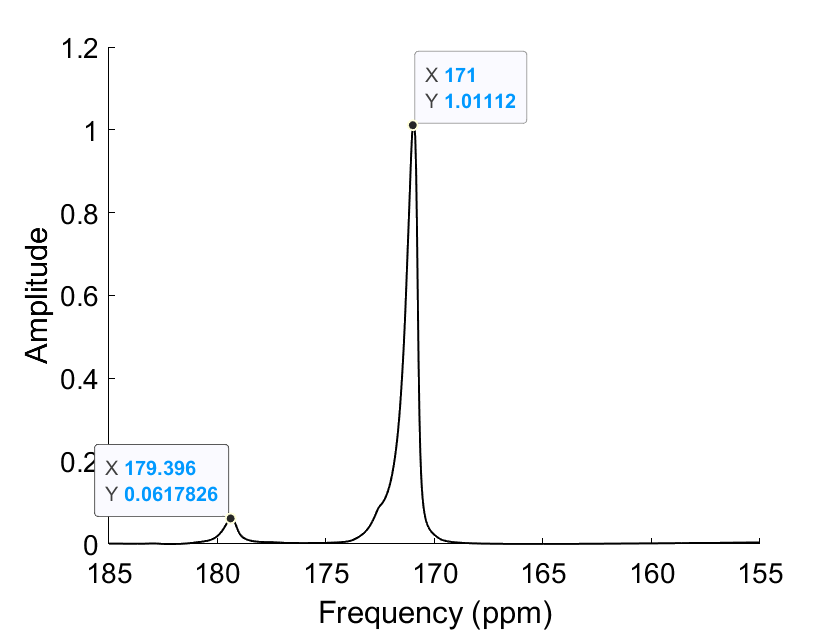
